# SARS-CoV-2 antibody prevalence by industry, workplace characteristics, and workplace infection prevention and control measures, North Carolina, 2021 to 2022

**DOI:** 10.1101/2024.03.06.24303821

**Authors:** Carolyn Gigot, Nora Pisanic, Kristoffer Spicer, Meghan F. Davis, Kate Kruczynski, Magdielis Gregory Rivera, Kirsten Koehler, D. J. Hall, Devon J. Hall, Christopher D. Heaney

## Abstract

**Background:** The COVID-19 pandemic has disproportionately affected workers in certain industries and occupations, and the workplace can be a high risk setting for SARS-CoV-2 transmission. In this study, we measured SARS-CoV-2 antibody prevalence and identified work-related risk factors in a population primarily working at industrial livestock operations.

**Methods:** We used a multiplex salivary SARS-CoV-2 IgG antibody assay to determine infection-induced antibody prevalence among 236 adult (≥18 years) North Carolina residents between February 2021 and August 2022. We used the National Institute for Occupational Safety and Health Industry and Occupation Computerized Coding System (NIOCCS) to classify employed participants’ industry and compared infection-induced IgG prevalence by participant industry and with the North Carolina general population. We also combined antibody results with reported SARS-CoV-2 molecular test positivity and vaccination history to identify evidence of prior infection. We used logistic regression to estimate odds ratios of prior infection by potential work-related risk factors, adjusting for industry and date.

**Results:** Most participants (55%) were infection-induced IgG positive, including 71% of animal slaughtering and processing industry workers, which is 1.5 to 4.3 times higher compared to the North Carolina general population, as well as higher than molecularly-confirmed cases and the only other serology study we identified of animal slaughtering and processing workers. Considering questionnaire results in addition to antibodies, the proportion of participants with evidence of prior infection increased slightly, to 61%, including 75% of animal slaughtering and processing workers. Participants with more than 1000 compared to 10 or fewer coworkers at their jobsite had higher odds of prior infection (adjusted odds ratio [aOR] 4.5, 95% confidence interval [CI] 1.0 to 21.0).

**Conclusions:** This study contributes evidence of the severe and disproportionate impacts of COVID-19 on animal processing and essential workers and workers in large congregate settings. We also demonstrate the utility of combining non-invasive biomarker and questionnaire data for the study of workplace exposures.

**Conflict of Interest:** The authors declare no potential conflicts of interest with respect to the research, authorship, and/or publication of this article.

**What’s important about this paper:** High numbers of COVID-19 outbreaks, cases, and deaths have been reported among livestock industry workers, including Black and Hispanic workers, in the United States. Little is known about SARS-CoV-2 infection as measured by antibody prevalence in this setting. Antibody-based estimates of SARS-CoV-2 infection can capture cases missed by SARS-CoV-2 molecular testing, which is important given limitations in worker access to molecular diagnostic testing. We observed high SARS-CoV-2 infection-induced IgG prevalence in animal slaughtering and processing industry workers (71%) between February 2021 and August 2022, which is 1.5 to 4.3 times higher compared to the North Carolina general population, as well as higher than molecularly-confirmed cases and the only serology study we identified of animal slaughtering and processing workers. We also found higher odds of SARS-CoV-2 infection among participants at worksites with larger compared to smaller numbers of employees.

## INTRODUCTION

COVID-19 continues to have serious adverse occupational health as well as public health impacts. Essential workers who provide critical services and functions, such as healthcare, social services, transportation, and food industry workers, were exempted from precautionary COVID-19 pandemic lockdown policies, generally cannot work from home, and have been harmed disproportionally (Carlsten et al. 2021; Mutambudzi et al. 2021; CDC 2024). COVID-19 outbreaks at United States (US) meat and poultry processing operations and long-term care facilities were reported early in spring 2020, and more than 12,000 workplace COVID-19 outbreaks were reported by 23 health departments during August-October 2021 (Luckhaupt et al. 2023). More than 59,000 worker COVID-19 cases were reported by five meat processing companies during the first year of the pandemic in the US (House Staff Memorandum 2021). Studies across the US have found higher COVID-19 mortality among particular occupational sectors, including farming, construction, production, transportation, and healthcare support, and higher mortality among Black and Hispanic workers (Hawkins et al. 2021; Billock et al. 2022; Cummings et al. 2022).

Understanding which job groups are at higher risk is foundational to target workplace safety interventions. The disproportionality of COVID-19 cases and deaths is connected to many interrelated factors. Co-morbidities, co-exposures, socioeconomic status, and limited access to paid sick leave or healthcare can increase workers’ susceptibility to disease and lead to more severe outcomes (Carlsten et al. 2021). Workplace characteristics and job tasks, including prolonged close contact with coworkers, clients, customers, or patients; insufficient ventilation; and lack of appropriate personal protective equipment (PPE) can increase workers’ exposures to SARS-CoV-2 (Carlsten et al. 2021). One study found a correlation between worker complaints to the US Occupational Safety and Health Administration (OSHA) related to COVID-19 and subsequent COVID-19 cases and deaths during January-September 2020, suggesting worker recognition of unsafe conditions (Hanage et al. 2020). Research has also associated a range of workplace SARS-CoV-2 infection prevention and control (IPC) measures–including surveillance, facilitating employees staying home when ill, improving ventilation, changes in work arrangement to reduce crowding, providing adequate PPE, and requiring universal masking–with reductions in COVID-19 cases (Ingram et al. 2021). However, most studies focused on hospital and nursing home settings, with fewer assessing IPC measures in other high-risk workplaces, including livestock agriculture (Ingram et al. 2021).

SARS-CoV-2 antibody data can provide information on cumulative incidence of infection and is not affected by limited access to diagnostic testing or limited reporting of at-home rapid SARS-CoV-2 antigen test results (Pisanic et al. 2020). Antibodies are usually measured in blood; however, gingival crevicular fluid, which is rich in blood-derived IgG antibodies, can be used as a viable alternative to blood (Brandtzaeg 2007). SARS-CoV-2 antibodies measured in oral fluid (hereafter, saliva) have been shown to identify prior exposure to or infection with SARS-CoV-2 among PCR-confirmed COVID-19 cases with high sensitivity and specificity, with comparatively noninvasive and convenient sample collection that may facilitate participation, particularly among vulnerable or hard-to-reach populations (Pisanic et al. 2023). Vaccines approved for use in the US elicit antibodies specific to the SARS-CoV-2 spike (S) protein.

SARS-CoV-2 seroprevalence surveys have identified higher risk occupational groups, including meatpacking and farm workers, prior to widespread COVID-19 vaccine availability, but most focused on healthcare-related occupations (Klein et al. 2022; Boucher et al. 2023). Meza *et al*. used serology and infection and vaccination history to identify higher SARS-CoV-2 infection risk for in-person workers and certain occupational groups, including farming, fishing, and forestry, in California during summer 2021; however, their sample did not allow for quantification of infection risk among meatpacking or livestock workers (Meza et al. 2023).

In this study, we used a multiplex salivary SARS-CoV-2 IgG assay to determine infection-induced antibody prevalence in a North Carolina study population primarily working at industrial livestock operations. We compared infection-induced IgG prevalence by participant industry sector and with the North Carolina general population. We also combined SARS-CoV-2 IgG prevalence with reported viral test and vaccination history to determine any evidence of prior infection, and investigated associations between infection and work characteristics, including industry, essential worker status, and workplace IPC measures.

## METHODS

### Study design and population

This study was designed and conducted in a collaboration between the Johns Hopkins Bloomberg School of Public Health (BSPH) and the Rural Empowerment Association for Community Help (REACH), a community organization based in Duplin County, North Carolina, as previously described (Gigot et al. 2023). In brief, we recruited households in North Carolina, predominantly near industrial livestock operations, using a snowball sampling approach. Eligibility criteria included at least one adult (≥18) household member participating in the study, ability to understand spoken English or Spanish, and access to a household phone or mobile device and refrigerator. The BSPH Institutional Review Board (IRB) approved this study (IRB00014420).

Participants completed a baseline questionnaire that assessed demographics, work and workplace characteristics, and COVID-19 symptoms, testing, and vaccination history during a phone or video call with a study team member, who recorded responses in the secure web application REDCap (Harris et al. 2009; Harris et al. 2019). During the same baseline phone or video call, participants collected a saliva sample by swabbing their gum line with the ORACOL+ Saliva Collection Device (Malvern Medical Developments, Worcester, UK) for 1-2 minutes, and were instructed to store samples in a refrigerator until pickup or direct shipping. Saliva samples were tested for SARS-CoV-2 IgG antibodies with an in-house multiplex bead-based assay using Luminex technology which has been described previously (infection-induced [nucleocapsid (N) and spike [S] sensitivity=97.6% and specificity=99.4%; infection- and/or vaccination-induced [S] sensitivity, 99.4%; specificity, 99.3%) (Heaney et al. 2021; Gigot et al. 2023; Pisanic et al. 2023).

We have previously reported on demographics and antibody results for all participants of any age and industry (including not employed) enrolled between February 2021 and July 2022 (Gigot et al. 2023). Study enrollment continued for one additional month. Here, we report results for adult (≥18 years) participants enrolled over the full study period (February 2021 to August 2022) by industry and workplace characteristics.

### Industry classification

We used the National Institute for Occupational Safety and Health (NIOSH) Industry and Occupation Computerized Coding System (NIOCCS, https://csams.cdc.gov/nioccs/) to identify employed participants’ industry from free text job title, job category, and industrial livestock operation (ILO) category (industrial hog operation, industrial poultry operation, meat processing, animal rendering) (CDC 2022a). Classifications were also checked for consistency with ILO operation name and with self-reported direct contact with live or dead animals at work, with review by REACH community organizers. NIOCCS provides detailed 2017 North American Industry Classification System (NAICS) codes, grouped into 21 major sectors. Subsectors with 10 participants or fewer were grouped by major sector; major sectors with 10 participants or fewer were combined into an “Other” category.

### Identification of North Carolina general population N seroprevalence

We used SeroHub (data version 3.1.0, last updated 8/16/2023) to identify 6 SARS-CoV-2 infection-induced (N) seroprevalence studies of the North Carolina general population, 2 of which were excluded because North Carolina-specific estimates could not be identified in study text or supplement (Freedman et al. 2022). We did not identify any additional studies through a PubMed search (Table S1).

### Evidence of prior SARS-CoV-2 infection classification

Vaccines approved for use in the US elicit antibody responses to the receptor-binding domain of the SARS-CoV-2 S protein. Thus, antibodies against SARS-CoV-2 S indicate vaccination, response to infection, or both vaccination and infection; antibodies against SARS-CoV-2 N protein indicate response to infection (Duarte et al. 2022). However, N antibodies generally persist for a shorter period of time compared to S (Dan et al. 2021; Dhakal et al. 2023). To accurately characterize participants who had SARS-CoV-2 S antibodies in response to infection (because they had not received any vaccination doses) and whose N antibodies had waned over longer time since infection, as well as participants who reported a COVID-19 diagnosis via viral test, we used S and N antibody results combined with participants’ history of vaccination and positive viral test to determine evidence of prior SARS-CoV-2 infection (Figure 1). Since infection with SARS-CoV-2 usually elicits antibody responses to N and to S, we only classified samples with antibodies to N and to S as indicative of prior infection, as described in Pisanic *et. al.*, 2023 (Pisanic et al. 2023). This approach achieves high sensitivity and specificity to accurately distinguish between those with versus without prior SARS-CoV-2 infection (Pisanic et al. 2023).

**Figure 1.**
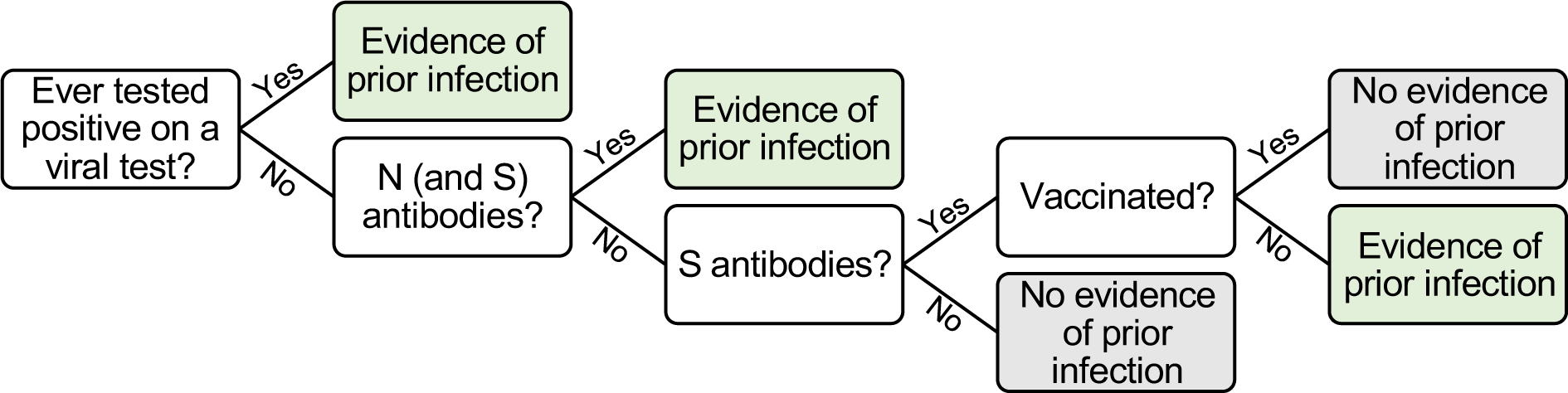
Conceptual decision tree for determining evidence of prior SARS-Cov-2 infection (green boxes) or no evidence of prior infection (grey boxes) using antibody (anti-N and anti-S antibodies) and questionnaire data (ever tested positive on a viral test, ever received any COVID-19 vaccine), based on Duarte et al. 2022

#### Statistical analysis

We tested for any differences in demographic characteristics and COVID-19 outcomes between industry groups using the Kruskal-Wallis rank sum test for continuous variables; Pearson’s Chi-squared test for categorical variables with all expected cell counts ≥5; and Fisher’s exact test for categorical variables with any expected cell count <5.

We used binomial regression with a logit link to calculate crude odds ratios (ORs) and 95% confidence intervals (95% CIs) comparing evidence of prior infection with SARS-CoV-2 between adult (≥18) participants in each industry compared to employed adult participants in all other industries, as well as between employed adult participants reporting each workplace characteristic or infection prevention and control (IPC) measures and those reporting absence of that characteristic or IPC measure. Where the characteristic or IPC measure question was categorical (e.g., number of other workers at worksite), we used the lowest level category as the reference group. We also calculated ORs for the association of industry with prior SARS-CoV-2 infection adjusted for sampling date and for sampling date and number of household occupants. For ORs of the association of workplace characteristics or IPC measures with infection, we adjusted for industry and for industry and sampling date. Number of household occupants was modelled as categorical (live alone; live with 1-2 cohabitants; live with >2 cohabitants) and sampling date was modelled as continuous (days from WHO COVID-19 pandemic declaration, March 11, 2020, (Ghebreyesus 2020) to sampling date). We used multiple imputation with chained equations to account for missing values in multivariable analyses (<1% missing overall). Using the *mice* package, we generated 5 imputed datasets using predictive mean matching for numeric data, logistic regression for binary data, and polytomous regression for categorical data, ran analyses with each imputed dataset, and pooled estimates, with total variance over repeated analyses computed using Rubin’s rules (Donald B. Rubin 1987; van Buuren and Groothuis-Oudshoorn 2011). All statistical analyses were completed in R 4.2.2 (R Core Team 2022). Because of the limitations of relying on a binary concept of statistical significance, we instead interpret results considering the magnitude, direction, and precision of effect estimates given our sample size and prior substantive knowledge (Amrhein et al. 2019).

## RESULTS

### Participant characteristics

Of 236 adult participants, 167 were employed: 57 in animal slaughtering and processing (North American Industry Classification System [NAICS] 31161, henceforth, animal processing), 20 in health care and social assistance (NAICS 62), 13 in animal production and aquaculture (NAICS 112, henceforth, animal production), and 77 in other industries, including 10 in administrative and support and waste management and remediation services, 10 in manufacturing (excluding animal processing), 8 in accommodation and food services, and 8 in retail trade (Table 1). Participants who were not employed were generally oldest (median 58 years), followed by animal production workers (median 46 years), health care and social assistance workers (median 38 years), participants employed in other industries (median 36 years), and animal processing workers (median 36 years). Gender distribution varied across industries, ranging from 15% female in animal production to 90% female in health care and social assistance. Most participants were Black (86% overall) or Hispanic/Latino (9% overall). Level of education varied by industry. Nearly half of health care and social assistance workers completed college, compared to zero animal production workers and 7% of animal processing workers. The number of household occupants also varied by industry: 32% of animal processing workers reported living with at least two other household members, compared to 8.3% of animal production workers and 7.4% of participants not employed. Most participants had some form of health insurance, although 11% overall were uninsured. Participants were enrolled between February 2021 and August 2022, and participation date varied by industry: health care and social assistance workers were generally enrolled earliest, followed by other industry workers, not employed, animal production workers, and animal processing workers.

**Table 1.**
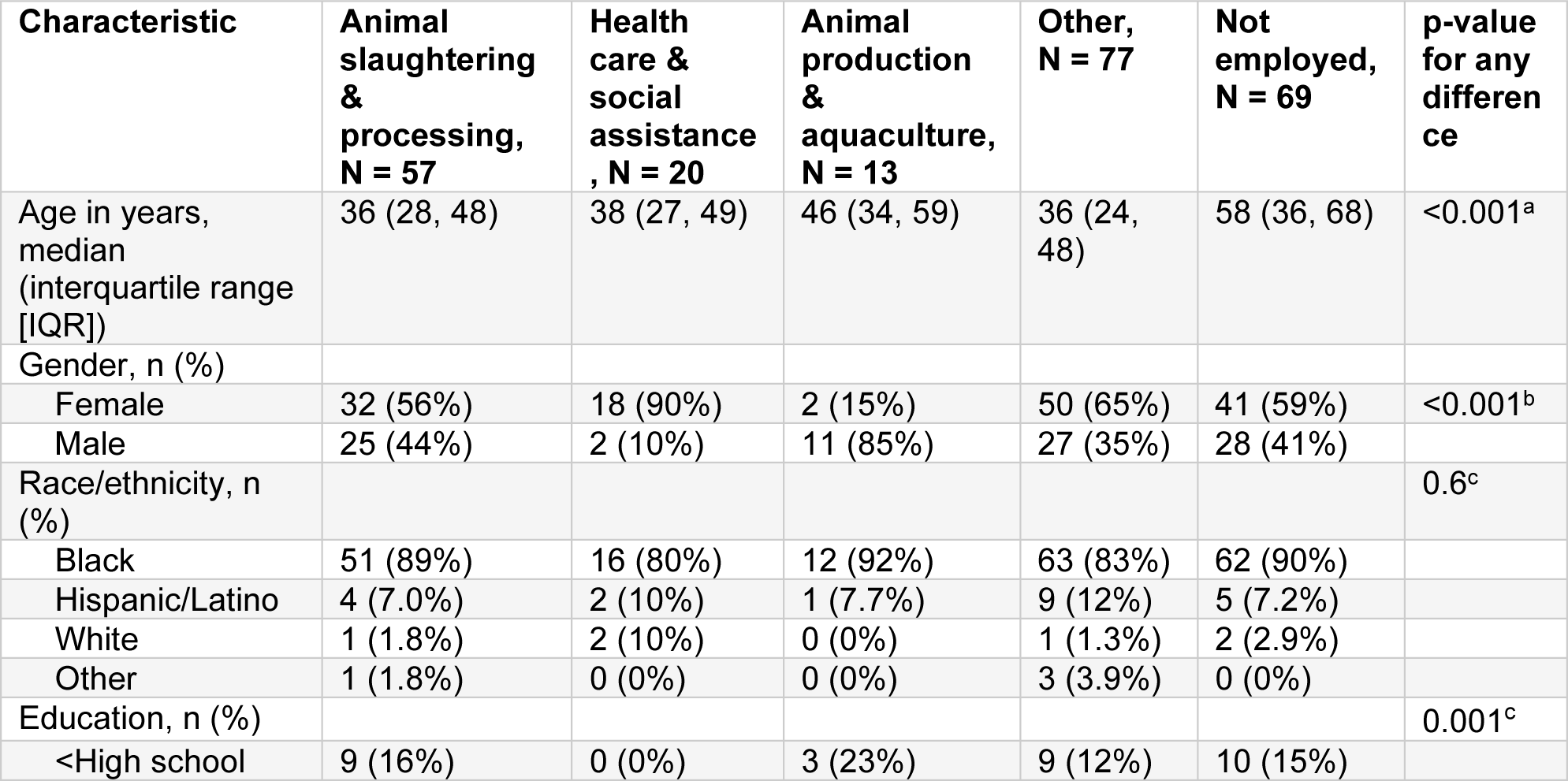
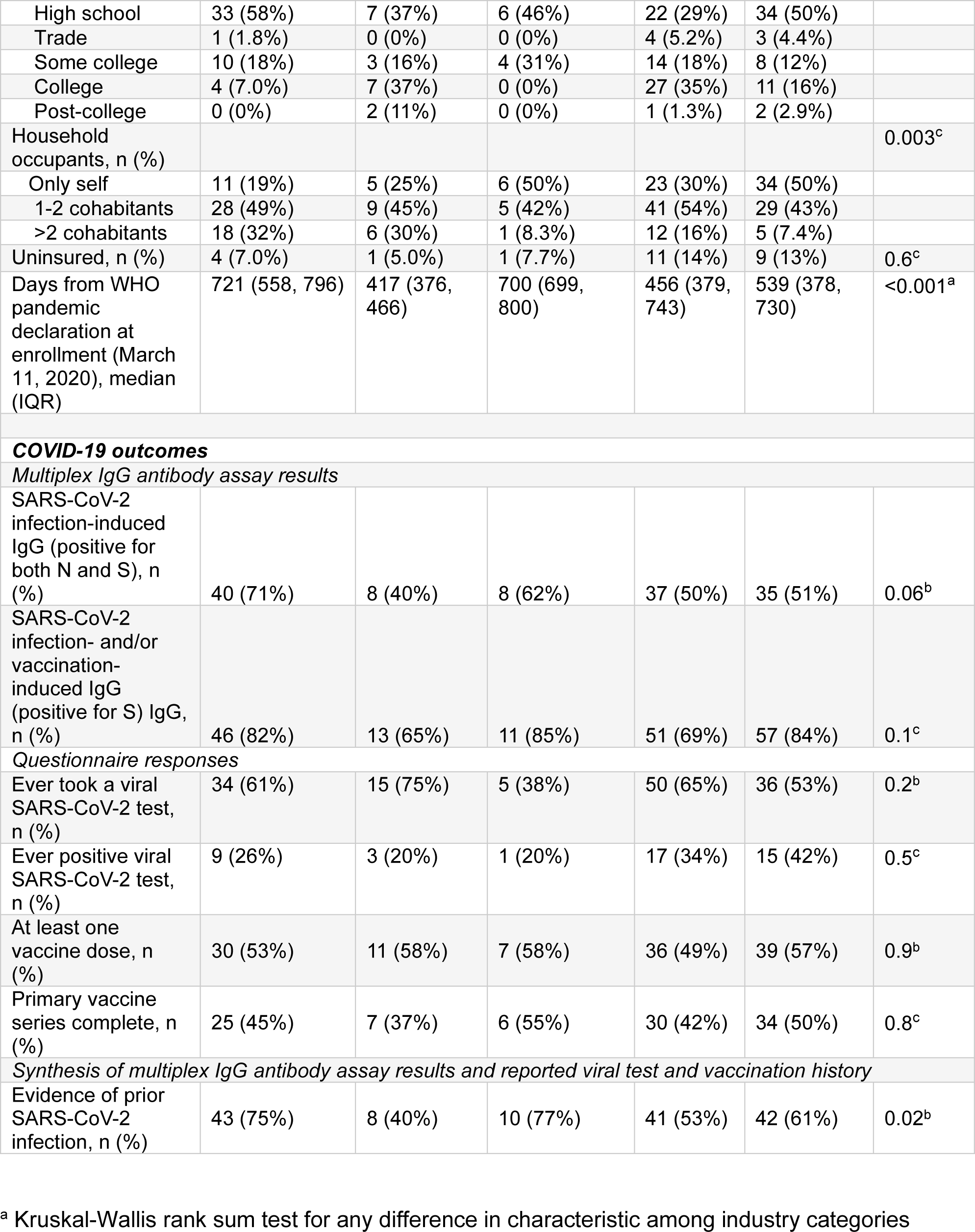
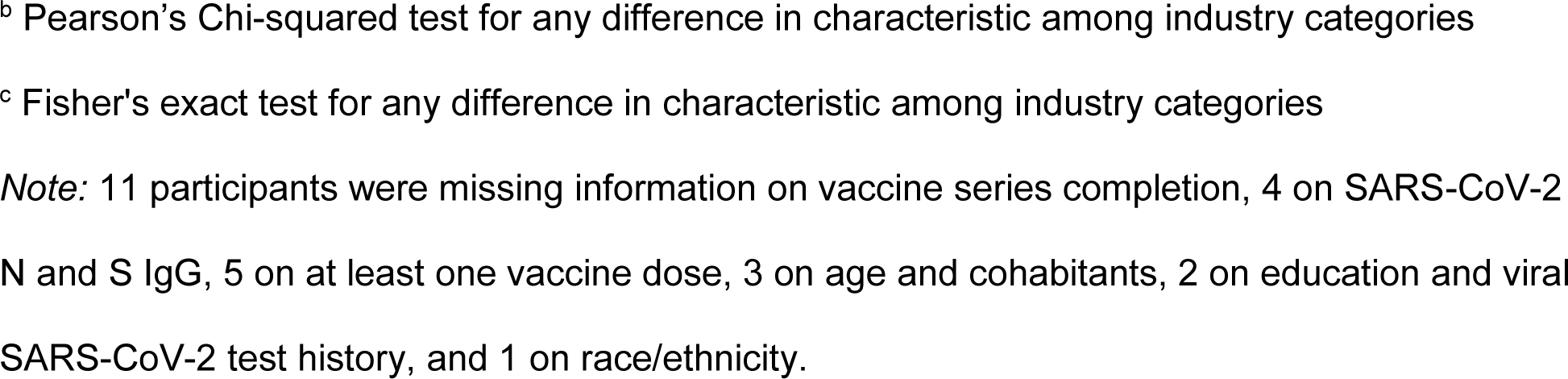
Adult (≥18) participant (N=236) characteristics by industry, North Carolina, 2021-2022.

| Characteristic | Animal slaughtering & processing, N = 57 | Health care & social assistance, N = 20 | Animal production & aquaculture, N = 13 | Other, N = 77 | Not employed, N = 69 | p-value for any difference |
| --- | --- | --- | --- | --- | --- | --- |
| Age in years, median (interquartile range [IQR]) | 36 (28, 48) | 38 (27, 49) | 46 (34, 59) | 36 (24, 48) | 58 (36, 68) | <0.001<sup>a</sup> |
| Gender, n (%) |  |  |  |  |  |  |
| Female | 32 (56%) | 18 (90%) | 2 (15%) | 50 (65%) | 41 (59%) | <0.001<sup>b</sup> |
| Male | 25 (44%) | 2 (10%) | 11 (85%) | 27 (35%) | 28 (41%) |  |
| Race/ethnicity, n (%) |  |  |  |  |  | 0.6<sup>c</sup> |
| Black | 51 (89%) | 16 (80%) | 12 (92%) | 63 (83%) | 62 (90%) |  |
| Hispanic/Latino | 4 (7.0%) | 2 (10%) | 1 (7.7%) | 9 (12%) | 5 (7.2%) |  |
| White | 1 (1.8%) | 2 (10%) | 0 (0%) | 1 (1.3%) | 2 (2.9%) |  |
| Other | 1 (1.8%) | 0 (0%) | 0 (0%) | 3 (3.9%) | 0 (0%) |  |
| Education, n (%) |  |  |  |  |  | 0.001<sup>c</sup> |
| <High school | 9 (16%) | 0 (0%) | 3 (23%) | 9 (12%) | 10 (15%) |  |
| High school | 33 (58%) | 7 (37%) | 6 (46%) | 22 (29%) | 34 (50%) |  |
| Trade | 1 (1.8%) | 0 (0%) | 0 (0%) | 4 (5.2%) | 3 (4.4%) |  |
| Some college | 10 (18%) | 3 (16%) | 4 (31%) | 14 (18%) | 8 (12%) |  |
| College | 4 (7.0%) | 7 (37%) | 0 (0%) | 27 (35%) | 11 (16%) |  |
| Post-college | 0 (0%) | 2 (11%) | 0 (0%) | 1 (1.3%) | 2 (2.9%) |  |
| Household occupants, n (%) |  |  |  |  |  | 0.003<sup>c</sup> |
| Only self | 11 (19%) | 5 (25%) | 6 (50%) | 23 (30%) | 34 (50%) |  |
| 1-2 cohabitants | 28 (49%) | 9 (45%) | 5 (42%) | 41 (54%) | 29 (43%) |  |
| >2 cohabitants | 18 (32%) | 6 (30%) | 1 (8.3%) | 12 (16%) | 5 (7.4%) |  |
| Uninsured, n (%) | 4 (7.0%) | 1 (5.0%) | 1 (7.7%) | 11 (14%) | 9 (13%) | 0.6<sup>c</sup> |
| Days from WHO pandemic declaration at enrollment (March 11, 2020), median (IQR) | 721 (558, 796) | 417 (376, 466) | 700 (699, 800) | 456 (379, 743) | 539 (378, 730) | <0.001<sup>a</sup> |
| **COVID-19 outcomes** |  |  |  |  |  |  |
| *Multiplex IgG antibody assay results* |  |  |  |  |  |  |
| SARS-CoV-2 infection-induced IgG (positive for both N and S), n (%) | 40 (71%) | 8 (40%) | 8 (62%) | 37 (50%) | 35 (51%) | 0.06<sup>b</sup> |
| SARS-CoV-2 infection- and/or vaccination-induced IgG (positive for S) IgG, n (%) | 46 (82%) | 13 (65%) | 11 (85%) | 51 (69%) | 57 (84%) | 0.1<sup>c</sup> |
| *Questionnaire responses* |  |  |  |  |  |  |
| Ever took a viral SARS-CoV-2 test, n (%) | 34 (61%) | 15 (75%) | 5 (38%) | 50 (65%) | 36 (53%) | 0.2<sup>b</sup> |
| Ever positive viral SARS-CoV-2 test, n (%) | 9 (26%) | 3 (20%) | 1 (20%) | 17 (34%) | 15 (42%) | 0.5<sup>c</sup> |
| At least one vaccine dose, n (%) | 30 (53%) | 11 (58%) | 7 (58%) | 36 (49%) | 39 (57%) | 0.9<sup>b</sup> |
| Primary vaccine series complete, n (%) | 25 (45%) | 7 (37%) | 6 (55%) | 30 (42%) | 34 (50%) | 0.8<sup>c</sup> |
| *Synthesis of multiplex IgG antibody assay results and reported viral test and vaccination history* |  |  |  |  |  |  |
| Evidence of prior SARS-CoV-2 infection, n (%) | 43 (75%) | 8 (40%) | 10 (77%) | 41 (53%) | 42 (61%) | 0.02<sup>b</sup> |

<sup>a</sup> Kruskal-Wallis rank sum test for any difference in characteristic among industry categories
<sup>b</sup> Pearson's Chi-squared test for any difference in characteristic among industry categories
<sup>c</sup> Fisher's exact test for any difference in characteristic among industry categories
*Note:* 11 participants were missing information on vaccine series completion, 4 on SARS-CoV-2 N and S IgG, 5 on at least one vaccine dose, 3 on age and cohabitants, 2 on education and viral SARS-CoV-2 test history, and 1 on race/ethnicity.

### SARS-CoV-2 infection-induced seroprevalence by industry and compared with the North Carolina general population

Most participants (55% overall) had infection-induced SARS-CoV-2 antibodies, including 71% of animal processing, 62% of animal production, 40% of health care and social assistance, 50% of other industry workers, and 51% of adult participants who were not employed (Table 1). Infection-induced seroprevalence was higher among animal processing industry workers in this study compared to all North Carolina general population infection-induced seroprevalence estimates identified (Figure 2). Infection-induced seroprevalence measured among animal processing industry workers ranged from 1.4 to 4.3 times North Carolina general population estimates during overlapping time periods identified in a literature search (Table S2). The overall study population seroprevalence was also higher than all North Carolina general population infection-induced seroprevalence estimates before February 2022 identified in a literature search.

**Figure 2.**
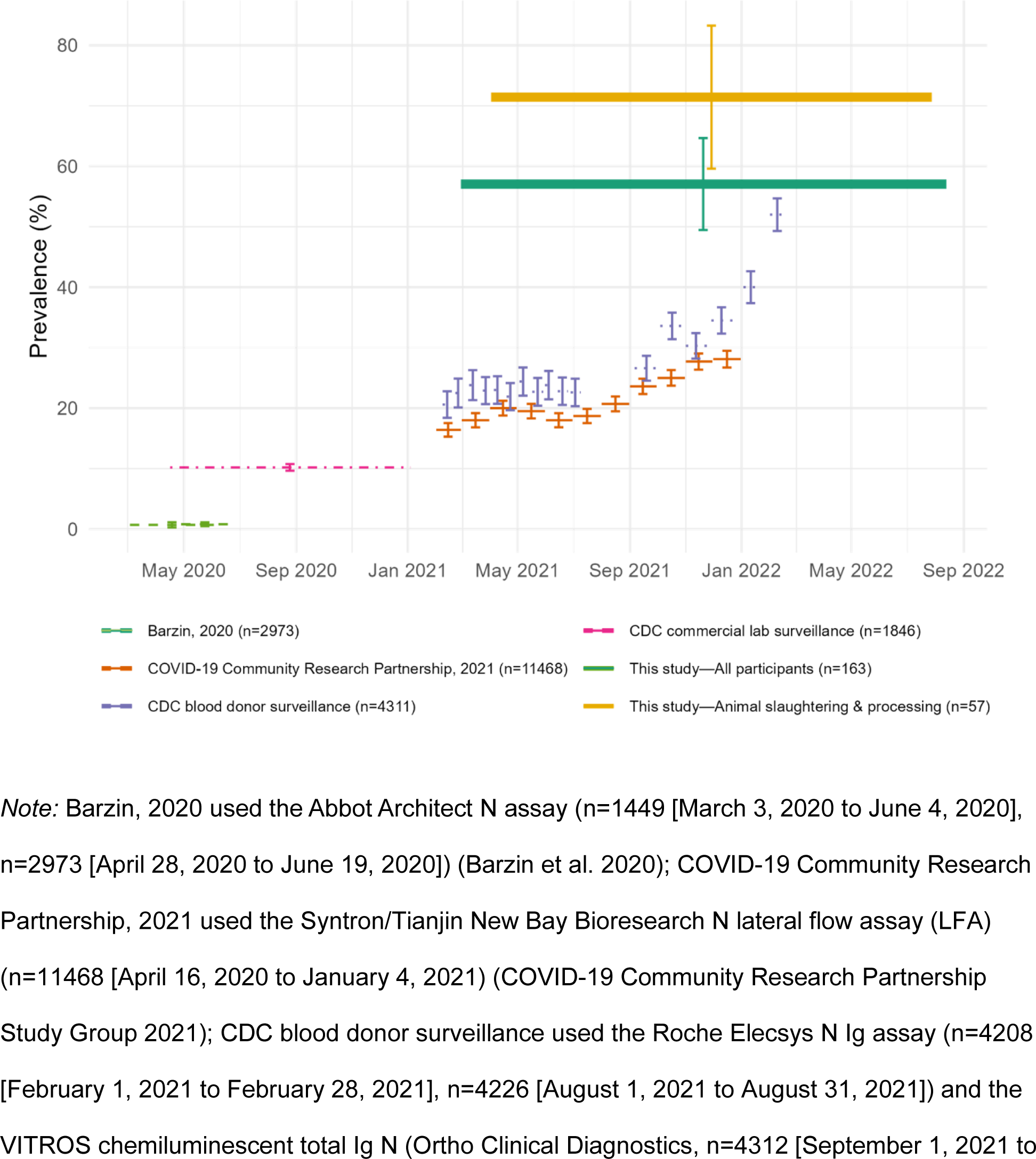
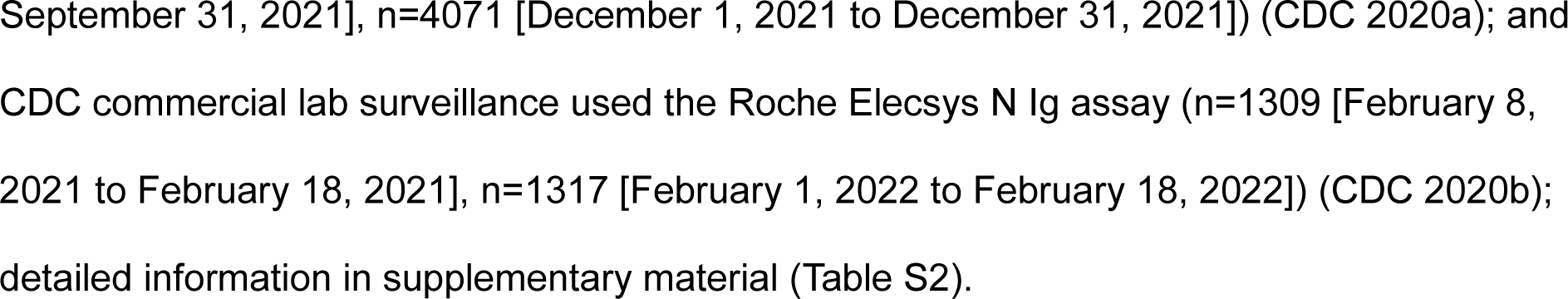
SARS-CoV-2 infection-induced antibody prevalence among animal slaughtering and processing industry workers (n=56) and all employed participants (n=163) in this study compared to North Carolina general population estimates, 2020-2022

### Evidence of prior SARS-CoV-2 infection by industry among employed participants

Most participants (77% overall) were positive for infection- and/or vaccination-induced (S) IgG, including 85% in animal production, 82% in animal processing, 40% in health care and social assistance, 69% in other industries, and 84% of adult participants who were not employed (Table 1). The proportion of participants who reported having taken a SARS-CoV-2 molecular or rapid antigen test before study participation and ever testing positive before study participation were similar across industries (60% and 32% overall, respectively). More than two thirds of participants (67%) had received at least one dose and 44% had completed the primary COVID-19 vaccination series.

Integrating multiplex antibody assay results and reported SARS-CoV-2 molecular or rapid antigen test results and vaccination history enabled the classification of 4 additional participants as having prior SARS-CoV-2 infection and resulted in classifications that largely corresponded with infection-induced antibody classification (Table S3). After combining antibody with self-reported viral test and vaccination history, most participants (61%) had evidence of prior SARS-CoV-2 infection, including 77% in animal production, 75% in animal processing, 40% in health care and social assistance, 53% in other industries, and 61% of adult participants who were not employed.

Using integrated multiplex antibody assay and questionnaire data, the odds of prior SARS-CoV-2 infection among animal processing workers was 2.7 (95% CI 1.3 to 5.4) times that of animal production, health care and social assistance, and other industry workers combined (Table 2). After adjusting for sampling date and household occupants, animal processing work was associated with 1.5 (95% CI 0.6 to 3.3) times the odds of SARS-CoV-2 infection. The odds of prior SARS-CoV-2 infection among health care and social assistance workers were 0.4 (95% CI 0.1 to 1.0) times those of participants employed in animal processing, animal production, and other industries. After adjusting for sampling date and household occupants, health care and social assistance work was associated with 0.5 (95% CI 0.2 to 1.6) times the odds of SARS-CoV-2 infection.

**Table 2.** Odds of prior SARS-CoV-2 infection among adult (≥18) employed participants in each industry sector compared to those in all others (N=167), North Carolina, 2021-2022.

| Industry sector | Prior infection/total, n (%) | Reference (employed in all other industry sectors) | OR (95% CI) | Model A aOR (95% CI) | Model B aOR (95% CI) |
| --- | --- | --- | --- | --- | --- |
| Animal production & aquaculture | 10/13 (77) | 92/154 (60) | 2.2 (0.6, 8.6) | 1.2 (0.3, 4.9) | 1.3 (0.3, 5.5) |
| Animal slaughtering & processing | 43/57 (75) | 59/110 (54) | 2.7 (1.3, 5.4) | 1.6 (0.7, 3.5) | 1.5 (0.6, 3.3) |
| Other | 41/77 (53) | 61/90 (68) | 0.6 (0.3, 1.1) | 0.9 (0.5, 1.9) | 1.0 (0.5, 2.1) |
| Health care & social assistance | 8/20 (40) | 94/147 (64) | 0.4 (0.1, 1.0) | 0.6 (0.2, 1.8) | 0.5 (0.2, 1.6) |

*Note:* OR = odds ratio; CI = confidence interval; aOR = adjusted odds ratio. Model A is adjusted for sampling date (modelled as continuous: days from WHO COVID-19 pandemic declaration, March 11, 2020 (Ghebreyesus 2020), to sampling date); Model B is adjusted for sampling date and number of household occupants (modelled as categorical: live alone, live with 1-2 cohabitants, live with >2 cohabitants). Two participants were missing information on the number of household occupants.

### Evidence of prior SARS-CoV-2 infection by workplace characteristics among employed participants

Across all industries, the odds of prior SARS-CoV-2 infection among participants identifying themselves as essential workers was 1.9 (95% CI 0.9 to 3.9) times the odds of employed participants not identifying themselves as essential workers (Table 3). After adjusting for industry, essential worker status was associated with 1.2 (95% CI 0.5 to 2.8) times the odds of infection. After adjusting for industry and sampling date, essential worker status was associated with 2.2 (95% CI 0.8 to 5.7) times the odds of infection (Figure 3). The odds of prior SARS-CoV-2 infection was 7.7 (95% CI 2.0 to 29.8) times among participants reporting more than 1000 versus 10 or fewer employees at their worksite. After adjusting for industry category, reporting more than 1000 compared to 10 or fewer employees at the worksite was associated with 6.4 (95% CI 1.4 to 28.8) times higher odds of SARS-CoV-2 infection. After adjusting for industry and sampling date, reporting more than 1000 compared to 10 or fewer employees at the worksite was associated with 4.5 (95% CI 1.0 to 21.0) times higher odds of SARS-CoV-2 infection.

**Figure 3.**
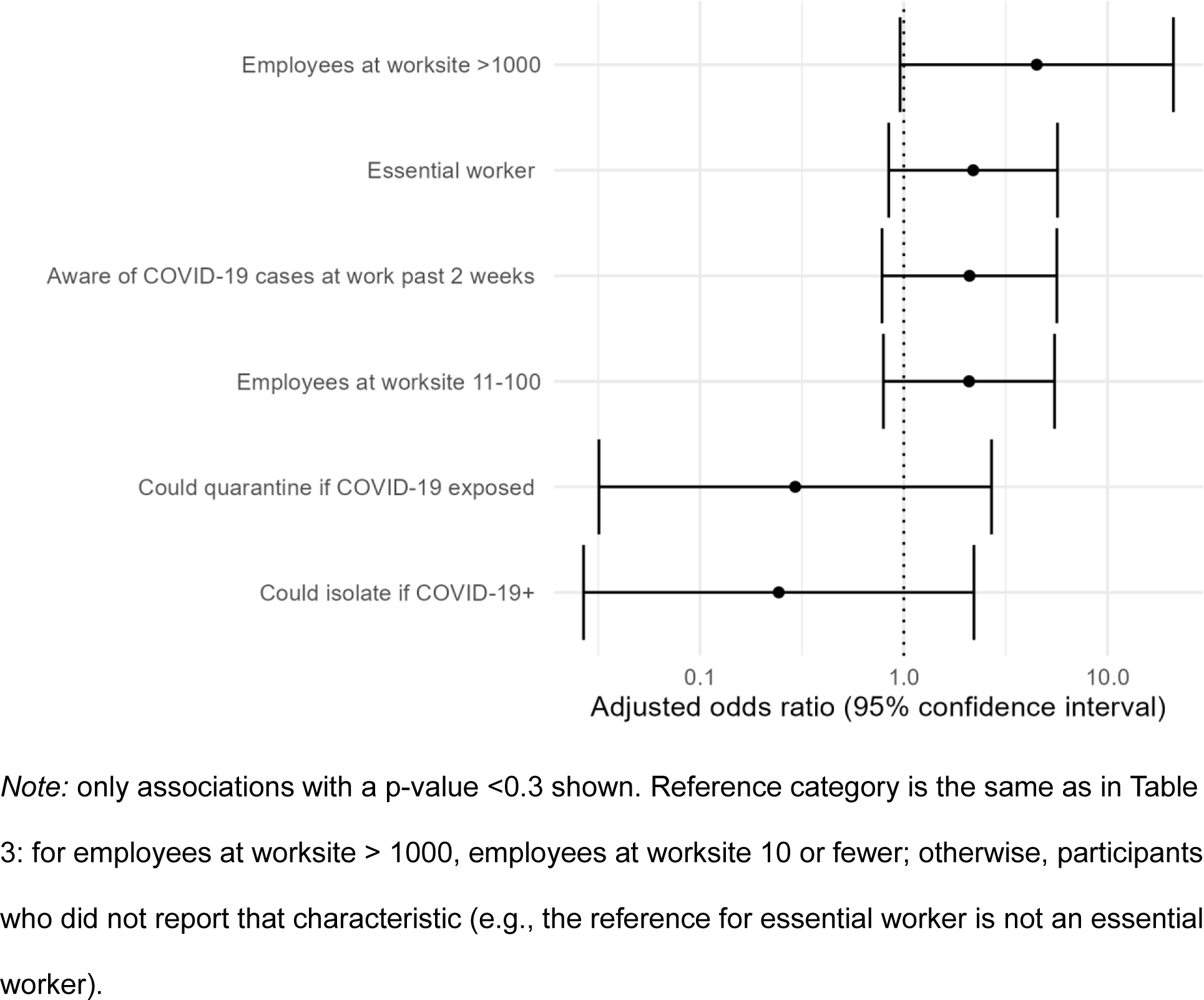
Adjusted odds (for industry sector and sampling date) of prior SARS-CoV-2 infection by workplace characteristics and infection prevention and control measures among adult (≥18) employed participants (N=167), North Carolina, 2021-2022

**Table 3.** Odds of prior SARS-CoV-2 infection by workplace characteristics and infection prevention and control measures among adult (≥18) employed participants (N=167), North Carolina, 2021-2022.

| Characteristic | Prior infection/total, n (%) | OR (95% CI) | Model A aOR (95% CI) | Model B aOR (95% CI) |
| --- | --- | --- | --- | --- |
| Essential worker |  |  |  |  |
| No | 20/41 (49) | - | - | - |
| Yes | 80/124 (65) | 1.9 (0.9, 3.9) | 1.2 (0.5, 2.8) | 2.2 (0.8, 5.7) |
| Worked in person past 2 weeks (at all) |  |  |  |  |
| No | 8/15 (53) | - | - | - |
| Yes | 94/152 (62) | 1.4 (0.5, 4.2) | 1.0 (0.3, 3.1) | 0.9 (0.3, 3.0) |
| Employees at worksite |  |  |  |  |
| 10 or fewer (reference) | 21/43 (49) | - | - | - |
| 11-100 | 32/48 (67) | 2.1 (0.9, 4.9) | 2.5 (1, 6.2) | 2.1 (0.8, 5.5) |
| 101-1000 | 24/43 (56) | 1.3 (0.6, 3.1) | 1.2 (0.5, 3.1) | 1.0 (0.4, 2.8) |
| >1000 | 22/25 (88) | 7.7 (2.0, 29.8) | 6.4 (1.4, 28.8) | 4.5 (1.0, 21.0) |
| Hours worked per week |  |  |  |  |
| <40 | 19/35 (54) | - | - | - |
| 40 | 31/52 (60) | 1.2 (0.5, 3.0) | 1.1 (0.4, 2.8) | 1.4 (0.5, 3.9) |
| >40 | 52/80 (65) | 1.6 (0.7, 3.5) | 0.9 (0.4, 2.3) | 1.3 (0.5, 3.3) |
| Aware of COVID-19 cases at work past 2 weeks |  |  |  |  |
| No | 12/23 (52) | - | - | - |
| Yes | 89/143 (62) | 1.5 (0.6, 3.7) | 1.6 (0.6, 4) | 2.1 (0.8, 5.6) |
| Able to maintain 6+ feet of distance |  |  |  |  |
| No | 21/39 (54) | - | - | - |
| Yes | 80/127 (63) | 1.5 (0.7, 3.0) | 1.4 (0.6, 3) | 1.2 (0.5, 2.8) |
| Could isolate if COVID-19+ |  |  |  |  |
| No | 9/10 (90) | - | - | - |
| Yes | 89/151 (59) | 0.2 (0.0, 1.3) | 0.2 (0.0, 1.5) | 0.2 (0.0, 2.2) |
| Could quarantine if COVID-19 exposed |  |  |  |  |
| No | 8/9 (89) | - | - | - |
| Yes | 90/152 (59) | 0.2 (0.0, 1.5) | 0.2 (0.0, 2.1) | 0.3 (0.0, 2.7) |
| **Infection prevention and control measures** |  |  |  |  |
| *Engineering controls* |  |  |  |  |
| Physical barriers between stations |  |  |  |  |
| No | 69/113 (61) | - | - | - |
| Yes | 33/54 (61) | 1.0 (0.5, 2.0) | 1.1 (0.5, 2.2) | 1.1 (0.5, 2.2) |
| Added hand washing stations |  |  |  |  |
| No | 31/53 (58) | - | - | - |
| Yes | 71/114 (62) | 1.2 (0.6, 2.3) | 1.0 (0.5, 2.1) | 1.4 (0.6, 3.0) |
| *Administrative controls* |  |  |  |  |
| Change in workplace sick leave |  |  |  |  |
| No | 77/119 (65) | - | - | - |
| Yes | 25/48 (52) | 0.6 (0.3, 1.2) | 0.8 (0.4, 1.6) | 1.0 (0.5, 2.2) |
| COVID-19 testing at work |  |  |  |  |
| No | 66/109 (61) | - | - | - |
| Yes | 36/58 (62) | 1.1 (0.6, 2.1) | 1.0 (0.5, 2.0) | 0.9 (0.4, 2.0) |
| *PPE* |  |  |  |  |
| Masks required |  |  |  |  |
| No | 24/37 (65) | - | - | - |
| Yes | 78/130 (60) | 0.8 (0.4, 1.7) | 0.8 (0.3, 1.7) | 1.2 (0.5, 2.9) |
| Employer provides face masks (any type) |  |  |  |  |
| No | 17/26 (65) | - | - | - |
| Yes | 85/141 (60) | 0.8 (0.3, 1.9) | 0.8 (0.3, 2.0) | 0.9 (0.3, 2.5) |
| N95/KN95/respirator |  |  |  |  |
| No | 81/132 (61) | - | - | - |
| Yes | 21/35 (60) | 0.9 (0.4, 2.0) | 1.0 (0.5, 2.3) | 0.7 (0.3, 1.8) |
| Surgical masks |  |  |  |  |
| No | 43/67 (64) | - | - | - |
| Yes | 59/100 (59) | 0.8 (0.4, 1.5) | 0.9 (0.4, 1.7) | 1.1 (0.5, 2.3) |
| Cloth masks |  |  |  |  |
| No | 76/126 (60) | - | - | - |
| Yes | 26/41 (63) | 1.1 (0.5, 2.4) | 1.1 (0.5, 2.4) | 1.0 (0.4, 2.3) |
| Employer provides face shields |  |  |  |  |
| No | 68/115 (59) | - | - | - |
| Yes | 34/52 (65) | 1.3 (0.7, 2.6) | 1.0 (0.5, 2.2) | 1.1 (0.5, 2.4) |
| Employer provides gloves |  |  |  |  |
| No | 46/81 (57) | - | - | - |
| Yes | 56/86 (65) | 1.4 (0.8, 2.7) | 1.0 (0.5, 2.1) | 1.1 (0.5, 2.4) |

*Note:* OR = odds ratio; CI = confidence interval; aOR = adjusted odds ratio. Model A is adjusted for industry (modelled as categorical: animal slaughtering and processing, health care and social assistance, animal production and aquaculture, other); model B is adjusted for industry and sampling date (modelled as continuous: days from WHO COVID-19 pandemic declaration, March 11, 2020 (Ghebreyesus 2020), to sampling date). Nine participants were missing information on number of employees at worksite; 8 on ability to isolate/quarantine; 3 on essential worker status; 2 on awareness of COVID-19 cases at work; and 1 on hours worked per week.

Adjusted for industry and sampling date, the odds of infection among participants reporting 11-100 employees at their worksite were 2.1 (95% CI 0.8 to 5.5) times the odds of those reporting 10 or fewer. Adjusted for industry and date, the odds of infection were 2.1 (95% CI 0.8 to 5.6) among participants reporting they were versus were not aware of COVID-19 cases at work during the past 2 weeks. Adjusted for industry and date, the odds of infection among participants reporting that they could isolate if testing positive without concern for their job were 0.2 (95% CI 0.0 to 2.2) those of participants reporting they could not. Similarly, the adjusted odds of infection among participants reporting that they could quarantine if COVID-19 exposed without concern for their job were 0.3 (95% CI 0.0 to 2.7) times those of participants reporting they could not.

### Evidence of prior SARS-CoV-2 infection by infection prevention and control (IPC) measures among employed participants

Adjusted for industry and date, the odds of SARS-CoV-2 infection among participants reporting that their employer added hand washing or sanitizing stations were 1.4 (95% CI 0.6 to 3.0) times those of participants reporting that they did not. Adjusted for industry and date, the odds of infection among participants reporting that their employer required masks at work were 1.2 (95% CI 0.5 to 2.9) times those of participants reporting they did not, while the odds of prior SARS-CoV-2 infection among participants reporting that their employer provided N95 or KN95 masks or respirators were 0.7 (95% CI 0.3 to 1.8) times those of participants reporting they did not (Table 3).

## DISCUSSION

The prevalence of SARS-CoV-2 infection-induced IgG prevalence among adults in our study population was 55%, almost double cumulative reported COVID-19 cases in North Carolina overall by the end of the study period (28.6% of the North Carolina population by August 2022) (US Census Bureau 2021; CDC 2023a) as well as higher than North Carolina\ general population infection-induced seroprevalence estimates from other studies (18% to 52% of the North Carolina general population) (Figure 1). Animal processing industry workers had the highest SARS-CoV-2 infection-induced IgG prevalence: 71%, 1.4 to 3.9-fold higher than the North Carolina general population infection-induced seroprevalence estimates during the same period, as well as about 6-fold higher than confirmed cases and higher than the only serology study we identified of workers in this industry (Klein et al. 2022). Data obtained from the five largest meatpacking companies in the US (which represent more than 60% of pork and 80% of beef production) found at least 59,000 confirmed COVID-19 cases during the first year of the pandemic (House Staff Memorandum 2021). Given the 519,450 workers employed in the animal slaughtering and processing industry in the US in 2021 (US Bureau of Labor Statistics 2022), at least 11.4% had confirmed COVID-19 by February 2021. Several factors could contribute to the 6-fold higher N seroprevalence we found, including infections that occurred after the first year of the pandemic, underreporting, asymptomatic and undiagnosed infections, and cases among workers in other companies and occupations in the animal slaughtering and processing industry. Given the generally shorter persistence of N antibodies, with an estimated half-life around 68 days (Dan et al. 2021), our results suggest continued high risk of SARS-CoV-2 infection for animal processing industry workers after the first year of the pandemic. We also found similarly high, or perhaps slightly higher infection-induced IgG seroprevalence compared to the only other animal slaughtering and processing industry worker seroprevalence study we identified, which found a seroprevalence of 64.6% among a population of North Carolina meatpacking workers in fall 2020 (Klein et al. 2022, Table S4). Our results are consistent with this high seroprevalence, but also suggest continued transmission and elevated risk during 2021 and 2022. Our results are also consistent with high numbers of reported COVID-19 outbreaks and deaths and concerns raised by workers and unions (UFCW 2020; Hawkins et al. 2021; Billock et al. 2022; Cummings et al. 2022; Luckhaupt et al. 2023). More than 95% of participants in our study were Black or Hispanic/Latino. The high rates of SARS-CoV-2 infection we observed are also consistent with higher seroprevalence among Black and Hispanic North Carolina residents described by Lopez et al (33.5% of Latinx and 17.1% of Black participants, compared to 10.5% of white participants) (Lopez et al. 2022) and higher seroprevalence among these groups across the US (40.2% of Hispanic and 32.5% of Black participants, compared to 27% of white participants) (Jones et al. 2022).

In addition to comparing SARS-CoV-2 infection-induced IgG prevalence by industry and with North Carolina general population seroprevalence, we integrated antibody test results with self-reported vaccination and diagnosis history to determine any evidence of prior infection, resulting in a slightly higher estimated prevalence of prior infection in our overall study population (61%), and the classification of 4 additional participants with prior SARS-CoV-2 infection compared to use of the IgG classification alone (Figure 1, Table S3). Using a multiplex immunoassay which defines evidence of prior SARS-CoV-2 infection based on N and S antibody results combined with vaccination and diagnosis history could improve classification of evidence of past infection (e.g., improve sensitivity to capture cases that would otherwise be missed), especially given the generally shorter persistence of N antibodies (Dan et al. 2021). This approach would accurately characterize participants who had S antibodies in response to infection and who had not received any vaccination doses whose N antibodies had waned over time since infection, as well as participants whose antibody response had waned below the limit of detection but who reported a prior COVID-19 diagnosis via molecular or rapid antigen SARS-CoV-2 virus test. This approach is subject to recall and social desirability bias, and participants with milder infections or more limited access to testing may not have been diagnosed with COVID-19. However, we believe participants were unlikely to not remember receiving a COVID-19 vaccination or a diagnosis of COVID-19.

We found higher odds of evidence of prior SARS-CoV-2 infection among animal processing workers compared to animal production, health care, and other industry workers combined, attenuated after adjusting for sampling date and household occupants. We also found lower odds of infection among health care and social assistance compared to animal processing, animal production, and other industry workers combined, attenuated after adjusting for sampling date and household occupants (Table 2). This is consistent with reports of high numbers of COVID-19 outbreaks, cases, and deaths among animal processing workers (Hawkins et al. 2021; House Staff Memorandum 2021; Billock et al. 2022; Cummings et al. 2022; Luckhaupt et al. 2023). Most studies of SARS-CoV-2 seroprevalence focus on healthcare workers, although some have found high seroprevalence among other essential workers, including farming workers (Boucher et al. 2023; Meza et al. 2023). Higher odds of SARS-CoV-2 infection among animal processing workers even in comparison to other largely high-risk worker groups (e.g., health care workers) underline the extremely high prevalence of COVID-19 infection we observed in this worker group. Attenuation after adjusting for other covariates also suggests that generally later enrollment dates and more household occupants account for some of the higher SARS-CoV-2 prevalence among animal processing workers. Research on outbreaks, severe outcomes, deaths, and seroprevalence have all found higher risks of COVID-19 among health care workers compared to the general population (Mutambudzi et al. 2021; Billock et al. 2022; Luckhaupt et al. 2023; Meza et al. 2023). However, a previous analysis of data from most participants in this study did find higher prevalence of SARS-CoV-2 infection-induced IgG among all industrial livestock operation workers and their household members compared to a healthcare worker cohort (prevalence ratio 2.5, 95% CI 1.8 to 3.3) (Gigot et al. 2023).The lower odds of SARS-CoV-2 we observed among health care and social assistance workers might be due to the high proportion of study participants working in other high-COVID-19-risk jobs (e.g., animal processing). Attenuation after adjusting for enrollment date also suggests that generally earlier enrollment dates account for some of the lower SARS-CoV-2 prevalence among health care and social assistance workers.

A previous analysis of most participants in this study did not identify increased infection-induced SARS-CoV-2 IgG prevalence in people working in meatpacking versus not (Gigot et al. 2023). The reference group in our previous analysis included participants who worked in the animal processing industry but not as meatpackers, e.g., in food service, maintenance, or sanitation jobs at large poultry or hog processing operations. We believe our industry classification based on all job and workplace questions better represents this highly exposed group compared to the meatpacking versus not meatpacking classification in the previous analysis. This analysis also uses a combination of SARS-CoV-2 IgG positivity, vaccination history, and COVID-19 diagnosis history to determine evidence of prior infection, rather than SARS-CoV-2 IgG alone, which could improve outcome classification, and includes additional adult participants recruited after the previous analysis, which could improve power.

Participants with more than 1000 employees at their worksite had higher adjusted odds of SARS-CoV-2 infection compared to participants with 10 or fewer (Table 3, Figure 3), which could be related to increased likelihood of contact with an infected coworker. Close contact with 10 or more coworkers has been associated with reported COVID-19 exposure at work among workers diagnosed with COVID-19 (Free et al. 2022). Many worksites with more than 1000 employees in this study were large pork processing facilities, with many other factors contributing to SARS-CoV-2 transmission risk—close proximity, long shifts, cold temperatures, low humidity, poor ventilation, and high levels of noise necessitating yelling to communicate (Taylor et al. 2020; Waltenburg et al. 2020; Klein et al. 2022). Essential workers in our study also had somewhat higher adjusted odds of SARS-CoV-2 infection, consistent with other studies showing that workers exempted from earlier lockdowns and generally unable to work from home were more likely to be exposed to infected coworkers, customers, and patients (Carlsten et al. 2021; Mutambudzi et al. 2021; Meza et al. 2023).

Participants who reported that they could isolate if testing positive or quarantine if exposed to someone with COVID-19 had somewhat lower adjusted odds of SARS-CoV-2 infection. Workplaces where employees take off when ill could be lower risk settings for SARS-CoV-2 transmission. Feeling able to take time off when ill could also reflect a constellation of other factors related to reduced COVID-19 risk, including workplace safety climate, workplace safety practices, and ability to take actions to reduce COVID-19 risk within and outside of the workplace.

The hierarchy of controls is a framework for prioritizing interventions to protect workers, with elimination of the hazard preferred, followed by substitution, engineering controls, administrative controls, and PPE (CDC 2023b). Fewer study participants reported engineering and administrative controls compared with PPE: 141 participants (84%) reported their employer provided face masks, compared to 54 (32%) reporting barriers between stations and 48 (29%) changes in workplace sick leave (Table 3, Table S5). Employers might preferentially implement simpler, lower cost interventions, especially given limited enforceable workplace standards. While OSHA’s PPE standard and general duty clause requiring employers provide workers a safe place of employment apply to COVID-19, OSHA issued a COVID-19 emergency temporary standard (ETS) only for healthcare workers, and withdrew non-recordkeeping portions of this ETS in December 2021 (Hanage et al. 2020; Michaels and Wagner 2020; OSHA 2021).

Comprehensive COVID-19 IPC measures (contact tracing and case isolation, facilitating smaller cohorts, viral testing, masking) have been associated with lower COVID-19 positivity in a variety of workplace settings (Ingram et al. 2021). Universal masking and physical barriers have been associated with reductions in COVID-19 incidence in meat processing facilities (Herstein et al. 2021). We did not find associations between any IPC measures we asked about and adjusted odds of SARS-CoV-2 infection, which could be because of our small sample size, changes over time, differences between industries, other factors with larger effects on COVID-19 risk, unmeasured or residual confounding, or some combination.

There are several limitations to this analysis. Participants were recruited via convenience and snowball sampling and might differ in several ways from the employed North Carolina population in general. Given the cross-sectional nature of our analysis, we were not able to establish the timing of industry employment and workplace characteristics before versus after SARS-CoV-2 infection. We sought to control for covariates related to both work and COVID-19 risk but cannot rule out unmeasured and residual confounding. Our study period (February 2021 to August 2022) included periods of dynamic rates of transmission of COVID-19 and changing public health and occupational health guidance, and our limited sample size prevented stratification by shorter time periods (CDC 2020a; CDC 2022b). We also did not ask about all COVID-19 infection prevention and control measures of interest, including employer support or requirements for worker vaccination. Our limited sample size prevented analysis by more detailed industry or occupational groups, which might have resulted in exposure misclassification or missed associations in smaller industry sectors or subsectors, and this is an important consideration for future work.

## CONCLUSIONS

This study adds to evidence of the severe, disproportionate effects of COVID-19 among livestock industry workers. Animal slaughtering and processing industry workers had higher rates of SARS-CoV-2 infection-induced antibody positivity compared with North Carolina general population estimates during the same period. Workers with more coworkers at their jobsite also had higher odds of evidence of prior SARS-CoV-2 infection compared to workers with fewer, adding to evidence of higher infection risk in large, congregate workplaces. The study also demonstrates the utility of participant self-collection of a less invasive salivary antibody assay and combining serology and questionnaire data for the study of occupational exposures.

## Acknowledgements

We thank the community organizers at REACH, especially Phyla Holmes, Margaret Carr, Clesha Hall, Unique Hall, Angela Matthews, Arika Miller, and Helen Santizo, as well as all participants, without whom this research would not have been possible. We acknowledge Dr. Pranay Randad for assistance with study design and launch. We acknowledge Dr. William A. Clarke, Caryn Kok, and Eric Xu for their work on SARS-CoV-2 antibody testing.

## Data availability

Deidentified data and code can be requested by email to the corresponding authors.

