## Supplemental Materials for "SARS-CoV-2 antibody prevalence by industry, workplace characteristics, and workplace infection prevention and control measures, North Carolina, 2021 to 2022"

***Table S1.*** *PubMed search strategy for North Carolina SARS-CoV-2 seroprevalence estimates*

| | |
|---|---|
| 1 | (Serology[Mesh] OR “Serologic Tests"[Mesh] OR Immunoglobulins[Mesh] OR “Seroepidemiologic Studies"[Majr] OR sero*[tiab] OR antibody[tiab] OR “anti-body”[tiab] OR antibodies[tiab]) |
| 2 | ("COVID-19"[Mesh] OR "SARS-CoV-2"[Mesh] OR “COVID-19”[tiab] OR coronavirus[tiab] OR “covid 2019”[tiab] OR “SARS-CoV-2”[tiab] OR “severe acute respiratory syndrome coronavirus 2”[tiab]) |
| 3 | "COVID-19 Serological Testing"[Mesh] |
| 4 | ("North Carolina"[Mesh] OR “North Carolina”[tiab] OR “Charlotte”[tiab] OR “Raleigh”[tiab] OR “Greensboro”[tiab] OR “Durham”[tiab] OR “Winson-Salem”[tiab] OR “Fayetteville”[tiab] OR “Cary”[tiab] OR “Wilmington”[tiab]) |
| 5 | (2019/12[Date - Publication]: 3000[Date - Publication]) |
| 6 | ((1 AND 2) OR 3) AND 4 AND 5 |

*Note:* 27 studies identified, none met all criteria or were not duplicates from SeroHub. Last date of search was October 16, 2023.

***Table S2.*** *SARS-CoV-2 infection-induced seroprevalence among the North Carolina general population by midpoint between collection start and end dates*

| Study | **Population description** | **Assay** | **Collection dates (year-month-day)** | **Sample size** | **Infection-induced seroprevalence, % (95% CI)** |
|---|---|---|---|---|---|
| Barzin, 2020 | Inpatients unrelated to COVID-19 | Abbot Architect N | 2020-03-03 to 2020-06-04 | 1449 | 0.7 |
| Barzin, 2020 | Asymptomatic people at NC outpatient clinics | Abbot Architect N | 2020-04-28 to 2020-06-19 | 2973 | 0.8 |
| COVID-19 Community Research Partnership Study Group, 2021 | Wake Forest Baptist Health and Atrium Health systems, general pop | Syntron/Tianjin New Bay Bioresearch N lateral flow assay (LFA) | 2020-04-16 to 2021-01-04 | 11468 | 10.2 |
| CDC commercial lab surveillance | NC commercial lab remnant specimens | Roche Elecsys N | 2021-02-08 to 2021-02-18 | 1309 | 20.6 (18.3,23) |
| CDC blood donor surveillance | Central and Western NC region blood donors | Roche Elecsys N | 2021-02-01 to 2021-02-28 | 4208 | 16.4 (14.6,18.5) |
| CDC commercial lab surveillance | NC commercial lab remnant specimens | Roche Elecsys N | 2021-02-22 to 2021-03-01 | 1172 | 22.5 (19.8,25) |

| | | | | | |
|---|---|---|---|---|---|
| CDC commercial lab surveillance | NC commercial lab remnant specimens | Roche Elecsys N | 2021-03-09 to 2021-03-18 | 1140 | 23.8 (21.1,26.6) |
| CDC blood donor surveillance | Central and Western NC region blood donors | Roche Elecsys N | 2021-03-01 to 2021-03-31 | 4072 | 18 (16.2,19.8) |
| CDC commercial lab surveillance | NC commercial lab remnant specimens | Roche Elecsys N | 2021-03-24 to 2021-03-31 | 1362 | 22.9 (20.5,25.6) |
| CDC commercial lab surveillance | NC commercial lab remnant specimens | Roche Elecsys N | 2021-04-05 to 2021-04-14 | 1304 | 23 (20.2,25.6) |
| CDC blood donor surveillance | Central and Western NC region blood donors | Roche Elecsys N | 2021-04-01 to 2021-04-30 | 4195 | 20 (17.9,22.3) |
| CDC commercial lab surveillance | NC commercial lab remnant specimens | Roche Elecsys N | 2021-04-19 to 2021-04-28 | 1303 | 21.9 (19.2,24.6) |
| CDC commercial lab surveillance | NC commercial lab remnant specimens | Roche Elecsys N | 2021-05-03 to 2021-05-12 | 1307 | 24.4 (22.1,27) |
| CDC blood donor surveillance | Central and Western NC region blood donors | Roche Elecsys N | 2021-05-01 to 2021-05-31 | 4159 | 19.5 (17.8,21.2) |
| CDC commercial lab surveillance | NC commercial lab remnant specimens | Roche Elecsys N | 2021-05-17 to 2021-05-29 | 1272 | 22.7 (20.1,25.6) |
| CDC commercial lab surveillance | NC commercial lab remnant specimens | Roche Elecsys N | 2021-05-31 to 2021-06-09 | 1268 | 23.8 (21.1,26.4) |
| CDC blood donor surveillance | Central and Western NC region blood donors | Roche Elecsys N | 2021-06-01 to 2021-06-30 | 4157 | 18 (16,20.1) |
| CDC commercial lab surveillance | NC commercial lab remnant specimens | Roche Elecsys N | 2021-06-14 to 2021-06-25 | 1315 | 22.8 (20.2,25.3) |
| CDC commercial lab surveillance | NC commercial lab remnant specimens | Roche Elecsys N | 2021-06-28 to 2021-07-09 | 1302 | 22.6 (20,25.2) |
| CDC blood donor surveillance | Central and Western NC region blood donors | Roche Elecsys N | 2021-07-01 to 2021-07-31 | 4168 | 18.7 (16.6,21.1) |
| CDC blood donor surveillance | Central and Western NC region blood donors | Roche Elecsys N | 2021-08-01 to 2021-08-31 | 4226 | 20.7 (18.6,22.9) |
| CDC blood donor surveillance | Central and Western NC region blood donors | VITROS chemiluminescent totat Ig N, Ortho Clinical Diagnostics | 2021-09-01 to 2021-09-30 | 4312 | 23.6 (22,25.3) |
| CDC commercial lab surveillance | NC commercial lab remnant specimens | Roche Elecsys N | 2021-09-06 to 2021-10-02 | 1767 | 26.6 (24.2,29.1) |

| | | | | | |
|---|---|---|---|---|---|
| CDC blood donor surveillance | Central and Western NC region blood donors | VITROS chemiluminescent totat Ig N, Ortho Clinical Diagnostics | 2021-10-01 to 2021-10-31 | 4330 | 25 (22.9,27.1) |
| CDC commercial lab surveillance | NC commercial lab remnant specimens | Roche Elecsys N | 2021-10-04 to 2021-10-30 | 1775 | 33.6 (30.7,36.5) |
| CDC commercial lab surveillance | NC commercial lab remnant specimens | Roche Elecsys N | 2021-11-01 to 2021-11-24 | 1846 | 30.3 (27.7,33) |
| CDC blood donor surveillance | Central and Western NC region blood donors | VITROS chemiluminescent totat Ig N, Ortho Clinical Diagnostics | 2021-11-01 to 2021-11-30 | 4311 | 27.7 (25.7,29.6) |
| CDC commercial lab surveillance | NC commercial lab remnant specimens | Roche Elecsys N | 2021-11-29 to 2021-12-22 | 1833 | 34.5 (32.1,37.2) |
| CDC blood donor surveillance | Central and Western NC region blood donors | VITROS chemiluminescent totat Ig N, Ortho Clinical Diagnostics | 2021-12-01 to 2021-12-31 | 4071 | 28.1 (26.1,30) |
| CDC commercial lab surveillance | NC commercial lab remnant specimens | Roche Elecsys N | 2022-01-03 to 2022-01-20 | 1329 | 40 (36.7,43.4) |
| CDC commercial lab surveillance | NC commercial lab remnant specimens | Roche Elecsys N | 2022-02-01 to 2022-02-18 | 1317 | 52 (48.9,55.2) |

*Note:* estimates overlapping with Animal Slaughtering and Processing industry enrollment (April 2, 2021 to July 28, 2022) are highlighted in grey. The minimum prevalence estimate overlapping with Animal Slaughtering and Processing industry enrollment (April 2, 2021 to July 28, 2022) was 18% and maximum 52%.

***Table S3.*** *Source of information (multiplex antibody and questionnaire data) for determining evidence of prior SARS-CoV-2 infection among adult (≥18) participants (N=236), concordant multiplex N/S and synthesis results highlighted green (n=220)*

| **Questionnaire responses** | **Multiplex salivary SARS-CoV-2 IgG antibody assay results** | **Synthesis of antibody assay results and reported viral test and vaccination history** | **n** |
|---|---|---|---|

| | | | | |
|---|---|---|---|---|
| Ever positive viral SARS-CoV-2 test | ≥1 vaccine dose | N/S | Evidence of prior infection | 26 |
| Ever positive viral SARS-CoV-2 test | ≥1 vaccine dose | S | Evidence of prior infection | 2 |
| Ever positive viral SARS-CoV-2 test | ≥1 vaccine dose | Unknown | Evidence of prior infection | 2 |
| Ever positive viral SARS-CoV-2 test | No vaccine | N/S | Evidence of prior infection | 9 |
| Ever positive viral SARS-CoV-2 test | No vaccine | none | Evidence of prior infection | 2 |
| Ever positive viral SARS-CoV-2 test | No vaccine | Unknown | Evidence of prior infection | 3 |
| Ever positive viral SARS-CoV-2 test | Unknown | N/S | Evidence of prior infection | 1 |
| No positive viral SARS-CoV-2 test | ≥1 vaccine dose | N/S | Evidence of prior infection | 26 |
| No positive viral SARS-CoV-2 test | No vaccine | N/S | Evidence of prior infection | 16 |
| No positive viral SARS-CoV-2 test | No vaccine | S | Evidence of prior infection | 1 |
| Unknown | ≥1 vaccine dose | N/S | Evidence of prior infection | 20 |
| Unknown | No vaccine | N/S | Evidence of prior infection | 29 |
| Unknown | No vaccine | S | Evidence of prior infection | 6 |
| Unknown | Unknown | N/S | Evidence of prior infection | 1 |
| No positive viral SARS-CoV-2 test | ≥1 vaccine dose | S | No evidence of prior infection | 27 |
| No positive viral SARS-CoV-2 test | ≥1 vaccine dose | none | No evidence of prior infection | 6 |
| No positive viral SARS-CoV-2 test | No vaccine | none | No evidence of prior infection | 16 |
| No positive viral SARS-CoV-2 test | Unknown | S | No evidence of prior infection | 1 |
| No positive viral SARS-CoV-2 test | Unknown | none | No evidence of prior infection | 2 |
| Unknown | ≥1 vaccine dose | S | No evidence of prior infection | 13 |
| Unknown | ≥1 vaccine dose | none | No evidence of prior infection | 1 |
| Unknown | No vaccine | none | No evidence of prior infection | 26 |

*Note:* of 236 adult participants, 128 were positive for any evidence of prior SARS-CoV-2 infection and N/S IgG; 92 were negative for any evidence of infection and N/S IgG; 11 were positive for evidence of infection and negative for N/S IgG; and 5 were positive for any evidence of infection and invalid for N/S IgG. Of 167 employed adult participants, 93 were positive for any evidence of prior SARS-CoV-2 infection and N/S IgG; 65 were negative for any evidence of infection and N/S IgG; 5 were positive for evidence of infection and negative for N/S IgG; and 4 were positive for any evidence of infection and invalid for N/S IgG.

***Table S4.*** *PubMed search strategy for US animal slaughtering and processing worker SARS-CoV-2 seroprevalence estimates*

| | |
|---|---|
| 1 | (Serology[Mesh] OR “Serologic Tests"[Mesh] OR Immunoglobulins[Mesh] OR “Seroepidemiologic Studies"[Majr] OR sero*[tiab] OR antibody[tiab] OR “anti-body”[tiab] OR antibodies[tiab]) |
| 2 | ("COVID-19"[Mesh] OR "SARS-CoV-2"[Mesh] OR “COVID-19”[tiab] OR “covid 2019”[tiab] OR “SARS-CoV-2”[tiab] OR “severe acute respiratory syndrome coronavirus 2”[tiab]) |
| 3 | "COVID-19 Serological Testing"[Mesh] |
| 4 | ("Meat-Packing Industry"[Mesh] OR “Poultry”[Mesh] OR “Cattle”[Mesh] OR “Swine”[Mesh] OR “meat pack*”[tiab] OR meatpack*[tiab] OR “meat process*”[tiab] OR slaughterer*[tiab] OR cutter*[tiab] OR meat[tiab] OR poultry[tiab] OR cattle[tiab] OR swine[tiab]) |
| 5 | ("Occupational Groups"[Mesh] OR "Work"[Mesh] OR “Workplace”[Mesh] OR work*[tiab] OR occupation*[tiab] OR industry*[tiab]) |
| 6 | (2019/12[Date - Publication]: 3000[Date - Publication]) |
| 7 | ((1 AND 2) OR 3) AND 4 AND 5 AND 6 |

*Note:* 20 studies identified, only 1 relevant and carried out in the US. Last date of search was December 14, 2023.

***Table S5.*** *Workplace characteristics and infection prevention and control measures by industry among adult (≥18) employed participants (N=167), North Carolina, 2021-2022*

| **Characteristic, n (%)** | **Animal slaughtering & processing, N = 57** | **Health care & social assistance, N = 20** | **Animal production & aquaculture, N = 13** | **Other, N = 77** | **p-value for any difference** |
|---|---|---|---|---|---|
| Essential worker | 56 (98%) | 16 (80%) | 13 (100%) | 39 (52%) | <0.001[a] |
| Worked in person past 2 weeks (at all) | 57 (100%) | 20 (100%) | 13 (100%) | 62 (81%) | <0.001[a] |
| Employees at worksite | | | | | |
| 10 or fewer | 5 (8.8%) | 7 (39%) | 9 (69%) | 22 (31%) | |

| | | | | | |
|---|---|---|---|---|---|
| 11-100 | 10 (18%) | 8 (44%) | 3 (23%) | 27 (38%) | |
| 101-1000 | 21 (37%) | 1 (5.6%) | 1 (7.7%) | 20 (28%) | |
| >1000 | 21 (37%) | 2 (11%) | 0 (0%) | 2 (2.8%) | |
| Hours worked per week | | | | | <0.001[a] |
| <40 | 1 (1.8%) | 5 (25%) | 3 (23%) | 26 (34%) | |
| 40 | 17 (30%) | 12 (60%) | 2 (15%) | 21 (27%) | |
| >40 | 39 (68%) | 3 (15%) | 8 (62%) | 30 (39%) | |
| Aware of COVID-19 cases at work past 2 weeks | 49 (86%) | 16 (80%) | 10 (83%) | 68 (88%) | 0.7[a] |
| Able to maintain 6+ feet of distance | 42 (75%) | 12 (60%) | 12 (92%) | 61 (79%) | 0.2[a] |
| Could isolate if COVID-19+ | 51 (96%) | 20 (100%) | 10 (77%) | 70 (93%) | 0.078[a] |
| Could quarantine if COVID-19 exposed | 49 (92%) | 20 (100%) | 11 (85%) | 72 (96%) | 0.2[a] |
| *Infection prevention and control measures* | | | | | |
| *Engineering controls* | | | | | |
| Physical barriers between stations | 18 (32%) | 6 (30%) | 2 (15%) | 28 (36%) | 0.5[a] |
| Added hand washing stations | 44 (77%) | 14 (70%) | 9 (69%) | 47 (61%) | 0.3[a] |
| *Administrative controls* | | | | | |
| Change in workplace sick leave | 12 (21%) | 12 (60%) | 1 (7.7%) | 23 (30%) | 0.004[a] |
| COVID-19 testing at work | 29 (51%) | 11 (55%) | 2 (15%) | 16 (21%) | <0.001[a] |
| Masks required | 50 (88%) | 18 (90%) | 7 (54%) | 55 (71%) | 0.011[a] |
| *PPE* | | | | | |
| Employer provides face masks (any type) | 53 (93%) | 18 (90%) | 7 (54%) | 63 (82%) | 0.007[a] |
| Employer provides N95/KN96/respirators | 9 (16%) | 7 (35%) | 4 (31%) | 13 (17%) | 0.2[a] |
| Employer provides surgical masks | 36 (63%) | 16 (80%) | 4 (31%) | 42 (55%) | 0.030[b] |
| Employer provides cloth masks | 28 (49%) | 4 (20%) | 0 (0%) | 33 (43%) | 0.003[b] |
| Employer provides face shields | 28 (49%) | 7 (35%) | 3 (23%) | 14 (18%) | 0.001[a] |
| Employer provides hand protection | 44 (77%) | 12 (60%) | 7 (54%) | 23 (30%) | <0.001[b] |

[a] Fisher's exact test for any difference in characteristic among industry categories

[b] Pearson's Chi-squared test for any difference in characteristic among industry categories
